## Supplementary tables and figures for "Longitudinal change in cognition in older adults in Uganda: a prospective population study"

Table 1. Detailed baseline characteristics by wave and sex

Table 2. Mean score and correlates of each cognition component

Table 3. Details of participants lost to follow up and died from Wave 1

Figure 1. Cognition scores over time by educational attainment, adjusted for age,  sex, socio-economic position, and BMI

Figure 2. Cognition scores over time by socio-economic position (SEP), adjusted for age, sex, educational attainment, and BMI

*Table 1. Detailed baseline characteristics by wave and sex*

| | | Wave 1 | | Wave 2 | | Wave 3 | |
|---|---|---|---|---|---|---|---|
| | | **Men** | **Women** | **Men** | **Women** | **Men** | **Women** |
| | | **n (%)** | **n (%)** | **n (%)** | **n (%)** | **n (%)** | **n (%)** |
| **Residence** | Rural | 106 (54) | 150 (48) | 1 (2) | 7 (7) | 55 (69) | 58 (60) |
| | Urban | 92 (47) | 161 (52) | 44 (98) | 74 (9) | 25 (31) | 38 (40) |
| **Age group** | 50-59 | 70 (35) | 108 (35) | 27 (60) | 53 (65) | 41 (51) | 40 (42) |
| | 60-69 | 51 (26) | 99 (32) | 15 (33) | 20 (25) | 26 (33) | 31 (32) |
| | 70-79 | 54 (27) | 73 (24) | 3 (7) | 6 (7) | 6 (8) | 18 (19) |
| | 80+ | 23 (12) | 31 (10) | 0 (0) | 2 (3) | 7 (9) | 7 (7) |
| **Education** | No formal education | 29 (15) | 89 (29) | 2 (4) | 13 (16) | 11 (14) | 24 (25) |
| | Less than primary | 82 (41) | 157 (51) | 22 (49) | 29 (36) | 38 (48) | 44 (46) |
| | Completed primary school | 26 (13) | 27 (9) | 9 (20) | 20 (25) | 17 (21) | 10 (10) |
| | More than primary | 61 (31) | 37 (12) | 12 (27) | 19 (24) | 14 (18) | 18 (19) |
| **Marital status** | Married/cohabiting | 119 (60) | 46 (15) | 29 (64) | 11 (14) | 58 (73) | 22 (23) |
| | Divorced/separated/never married | 40 (20) | 65 (21) | 13 (29) | 21 (26) | 14 (18) | 29 (31) |
| | Widowed | 39 (20) | 200 (64) | 3 (7) | 49 (61) | 8 (10) | 44 (46) |
| **Socio-economic position** | 1 (Lowest) | 48 (25) | 70 (23) | 5 (11) | 16 (20) | 12 (15) | 22 (23) |
| | 2 | 41 (21) | 69 (23) | 11 (24) | 21 (26) | 17 (22) | 14 (15) |
| | 3 | 33 (17) | 65 (21) | 17 (38) | 19 (24) | 21 (27) | 18 (19) |
| | 4 | 43 (22) | 62 (20) | 11 (24) | 18 (22) | 13 (17) | 23 (25) |
| | 5 (Highest) | 31 (16) | 39 (13) | 1 (2) | 6 (8) | 16 (20) | 17 (18) |
| **Tobacco use** | Never used tobacco | 89 (45) | 253 (82) | 25 (56) | 67 (83) | 39 (49) | 79 (82) |
| | Current tobacco use | 52 (26) | 37 (12) | 5 (11) | 6 (7) | 17 (21) | 11 (12) |

| | | | | | | | |
|---|---|---|---|---|---|---|---|
| | Previous tobacco use | 57 (29) | 20 (7) | 15 (33) | 8 (10) | 24 (30) | 6 (6) |
| **Alcohol use** | No | 37 (19) | 89 (29) | 4 (9) | 28 (35) | 22 (28) | 37 (39) |
| | Yes | 161 (81) | 221 (71) | 41 (91) | 53 (65) | 58 (73) | 59 (62) |
| **BMI (kg/m$^2$)** | <18.5 | 36 (18) | 35 (11) | 11 (24) | 11 (14) | 14 (18) | 8 (8) |
| | 18.5- | 133 (67) | 161 (52) | 30 (67) | 49 (61) | 58 (73) | 55 (57) |
| | 25- | 19 (10) | 66 (21) | 4 (9) | 12 (15) | 7 (9) | 19 (20) |
| | 30+ | 10 (5) | 49 (16) | 0 (0) | 9 (11) | 1 (1) | 14 (15) |
| **Hypertension[1]** | No hypertension | 114 (58) | 153 (49) | 30 (67) | 48 (59) | 51 (64) | 49 (51) |
| | Hypertension | 84 (42) | 158 (51) | 15 (33) | 33 (41) | 29 (36) | 47 (49) |
| **Diabetes[2]** | No | 191 (97) | 290 (94) | 45 (100) | 79 (98) | 77 (96) | 89 (93) |
| | Yes | 7 (4) | 19 (6) | 0 (0) | 2 (3) | 3 (4) | 7 (7) |
| **Stroke[3]** | No | 188 (95) | 291 (94) | 42 (93) | 79 (98) | 79 (99) | 92 (96) |
| | Yes | 10 (5) | 20 (6) | 3 (7) | 2 (3) | 1 (1) | 4 (4) |
| **Angina[3]** | No angina | 148 (75) | 230 (74) | 36 (80) | 62 (77) | 67 (84) | 81 (84) |
| | Angina | 49 (25) | 81 (26) | 9 (20) | 19 (24) | 13 (16) | 15 (16) |
| **HIV** | HIV negative | 113 (57) | 197 (63) | 2 (4) | 11 (14) | 0 | 0 |
| | HIV positive | 85 (43) | 114 (37) | 43 (96) | 70 (86) | 79 (100) | 92 (100) |

1. Defined as systolic blood pressure≥140, diastolic blood pressure≥90, or self-report of use of antihypertensive medication
2. Defined as clinician diagnosis of diabetes
3. Defined as clinician diagnosis or a suggestive history (hemisensory loss or hemiparesis lasting >24 hours for stroke; chest pain on exertion for angina)

*Table 2. Mean score and correlates of each cognition component*

| | | Recall | | | Digit span | | | Verbal fluency | | |
|---|---|---|---|---|---|---|---|---|---|---|
| | | **Mean cognition score** | **Linear regression coefficient[1] (95% CI)** | **p-value** | **Mean cognition score** | **Linear regression coefficient[1] (95% CI)** | **p-value** | **Mean cognition score** | **Linear regression coefficient[1] (95% CI)** | **p-value** |
| **Wave at recruitment** | 1 | 0.18 | Ref | <0.0001 | 0.008 | Ref | 0.003 | 0.05 | Ref | <0.0001 |
| | 2 | -0.27 | -0.74 (-0.94;-0.54) | | 0.006 | -0.34 (-0.53;-0.14) | | -0.29 | -0.49 (-0.69;-0.29) | |
| | 3 | -0.17 | -0.46 (-0.62;-0.30) | | 0.06 | -0.05 (-0.21;0.11) | | 0.31 | 0.15 (-0.02;0.31) | |
| **Residence** | Urban | -0.03 | Ref | 0.30 | -0.12 | Ref | 0.005 | 0.25 | Ref | 0.55 |
| | Rural | 0.09 | 0.07 (-0.07;0.22) | | 0.14 | 0.20 (0.06;0.34) | | -0.07 | -0.04 (-0.19;0.10) | |
| **Sex** | Male | -0.07 | Ref | 0.02 | 0.17 | Ref | 0.04 | 0.11 | Ref | 0.004 |
| | Female | 0.10 | 0.19 (0.03;0.36) | | -0.08 | -0.17 (-0.34;-0.01) | | 0.009 | -0.24 (-0.41;-0.07) | |
| **Age group** | 50-59 | 0.27 | Ref | <0.0001 | 0.30 | Ref | <0.0001 | 0.19 | Ref | <0.0001 |
| | 60-69 | 0.04 | -0.26 (-0.41;-0.11) | | -0.04 | -0.32 (-0.48;-0.17) | | 0.02 | -0.14 (-0.29;0.02) | |
| | 70-79 | -0.29 | -0.66 (-0.84;-0.48) | | -0.25 | -0.46 (-0.64;-0.28) | | -0.16 | -0.31 (-0.49;-0.13) | |
| | 80+ | -0.69 | -1.03 (-1.27;-0.79) | | -0.63 | -0.78 (-1.03;-0.54) | | -0.54 | -0.69 (-0.94;-0.44) | |
| **Education** | No formal education | -0.25 | Ref | 0.08 | -0.63 | Ref | <0.0001 | -0.35 | Ref | 0.005 |
| | Any formal education | 0.06 | 0.15 (-0.02;0.32) | | 0.16 | 0.56 (0.39;0.73) | | 0.08 | 0.24 (0.07;0.41) | |
| **Marital status** | Married | 0.01 | Ref | 0.69 | 0.19 | Ref | 0.35 | 0.16 | Ref | 0.71 |
| | Not married | -0.004 | -0.03 (-0.19;0.13) | | -0.11 | -0.07 (-0.23;0.08) | | -0.09 | -0.03 (-0.19;0.13) | |

| | | | | | | | | | | |
|---|---|---|---|---|---|---|---|---|---|---|
| **SEP** | 1 (poorest) | -0.31 | Ref | <0.0001 | -0.37 | Ref | 0.002 | -0.14 | Ref | 0.14 |
| | 2 | 0.04 | 0.38 (0.19;0.57) | | 0.001 | 0.31 (0.12;0.51) | | -0.04 | 0.11 (-0.09;0.30) | |
| | 3 | 0.001 | 0.26 (0.06;0.46) | | 0.05 | 0.22 (0.02;0.42) | | 0.14 | 0.2 (-0.01;0.4) | |
| | 4 | 0.21 | 0.42 (0.22;0.63) | | 0.27 | 0.4 (0.19;0.6) | | 0.12 | 0.2 (-0.01;0.41) | |
| | 5 (richest) | 0.05 | 0.56 (0.32;0.80) | | 0.06 | 0.33 (0.09;0.57) | | -0.05 | 0.29 (0.05;0.53) | |
| **Tobacco use** | Current | -0.14 | Ref | 0.66 | -0.16 | Ref | 0.98 | -0.07 | Ref | 0.86 |
| | Previous | -0.05 | -0.03 (-0.26;0.20) | | 0.02 | -0.01 (-0.23;0.22) | | 0.11 | 0.06 (-0.17;0.30) | |
| | Never | 0.04 | -0.08 (-0.27;0.11) | | 0.03 | 0.01 (-0.18;0.2) | | -0.006 | 0.02 (-0.17;0.22) | |
| **Alcohol** | No alcohol | 0.001 | Ref | 0.99 | 0.008 | Ref | 0.31 | -0.008 | Ref | 0.24 |
| | Alcohol | 0.004 | 0.06 (-0.09;0.21) | | -0.012 | 0.08 (-0.07;0.23) | | 0.03 | 0.09 (-0.06;0.24) | |
| **BMI** | <18.5 | -0.38 | -0.27 (-0.46;-0.08) | 0.08 | -0.26 | -0.13 (-0.32;0.06) | 0.16 | -0.22 | -0.13 (-0.32;0.07) | 0.44 |
| | 18.5- | 0.01 | Ref | | 0.04 | Ref | | 0.047 | Ref | |
| | 25- | 0.27 | 0.12 (-0.06;0.31) | | 0.17 | 0.08 (-0.11;0.26) | | 0.07 | 0.01 (-0.18;0.20) | |
| | 30+ | 0.03 | -0.1 (-0.33;0.13) | | -0.17 | -0.16 (-0.38;0.07) | | -0.08 | -0.12 (-0.35;0.11) | |
| **HTN** | No hypertension | -0.02 | Ref | 0.73 | 0.02 | Ref | 0.89 | -0.014 | Ref | 0.32 |
| | Hypertension | 0.02 | 0.02 (-0.11;0.16) | | -0.03 | -0.01 (-0.14;0.13) | | 0.018 | 0.07 (-0.07;0.21) | |
| **Diabetes** | No diabetes | -0.01 | Ref | 0.88 | -0.01 | Ref | 0.82 | -0.007 | Ref | 0.73 |
| | Diabetes | 0.27 | 0.02 (-0.29;0.34) | | 0.26 | -0.04 (-0.35;0.27) | | 0.17 | -0.06 (-0.37;0.26) | |
| **Stroke** | No stroke | -0.008 | Ref | 0.53 | -0.004 | Ref | 0.4 | -0.003 | Ref | 0.73 |
| | Stroke | 0.16 | 0.09 (-0.2;0.38) | | 0.09 | 0.12 (-0.17;0.42) | | 0.06 | 0.05 (-0.25;0.35) | |
| **Angina** | No angina | 0.03 | Ref | | 0.003 | Ref | | -0.02 | Ref | |

| | | | | | | | | | | |
|---|---|---|---|---|---|---|---|---|---|---|
| | Angina | -0.09 | -0.1 (-0.26;0.05) | 0.17 | 0.0001 | 0.06 (-0.1;0.21) | 0.47 | 0.08 | 0.15 (-0.01;0.30) | 0.06 |
| **HIV** | No HIV | 0.03 | Ref | 0.67 | -0.09 | Ref | 0.32 | -0.02 | Ref | 0.26 |
| | HIV | 0.05 | 0.04 (-0.14;0.21) | | 0.09 | -0.09 (-0.26;0.09) | | 0.11 | -0.10 (-0.28;0.08) | |

1. Multivariable linear regression, adjusted for sex, age group, residence, marital status, education, socioeconomic position, tobacco use, and BMI

*Table 3. Number from Wave 1 lost to follow up and died*

| | | Not lost to follow up (%) | Lost to follow up (%) |
|---|---|---|---|
| | **Total** | 378 (74) | 131 (26) |
| **Sex** | Male | 143 (38) | 55 (42) |
| | Female | 235 (62) | 76 (58) |
| **Residence** | Rural | 224 (59) | 32 (24) |
| | Urban | 154 (41) | 99 (76) |
| **Age group** | 50-59 | 136 (36) | 42 (32) |
| | 60-69 | 105 (28) | 45 (34) |
| | 70-79 | 94 (25) | 33 (25) |
| | 80+ | 43 (11) | 11 (8) |
| **Educational attainment**[1] | No formal education | 98 (26) | 20 (15) |
| | Any formal education | 279 (74) | 111 (85) |
| **Marital status** | Married/ cohabiting | 127 (34) | 38 (29) |
| | Not married | 251 (66) | 93 (71) |
| **Socio-economic position** | 1 (Lowest) | 87 (24) | 31 (24) |
| | 2 | 79 (21) | 31 (24) |
| | 3 | 78 (21) | 20 (15) |
| | 4 | 81 (22) | 24 (19) |
| | 5 (Highest) | 46 (12) | 24 (19) |
| **Stroke** | No | 12 (3) | 4 (3) |
| | Yes | 366 (97) | 127 (97) |

| | | | |
|---|---|---|---|
| **Angina** | No angina | 282 (75) | 96 (73) |
| | Angina | 95 (25) | 35 (27) |
| **HIV** | HIV negative | 238 (63) | 72 (55) |
| | HIV positive | 140 (37) | 59 (45) |
| **Cognition score groups** | 5 (highest) | 78 (21) | 24 (19) |
| | 4 | 71 (19) | 29 (22) |
| | 3 | 65 (17) | 26 (20) |
| | 2 | 78 (21) | 25 (19) |
| | 1 (lowest) | 86 (23) | 26 (20) |

1. Missing data (Educational attainment, angina, cognition score, CV disease: missing data for 1 participant; Socio-economic position: missing data for 8 participants)

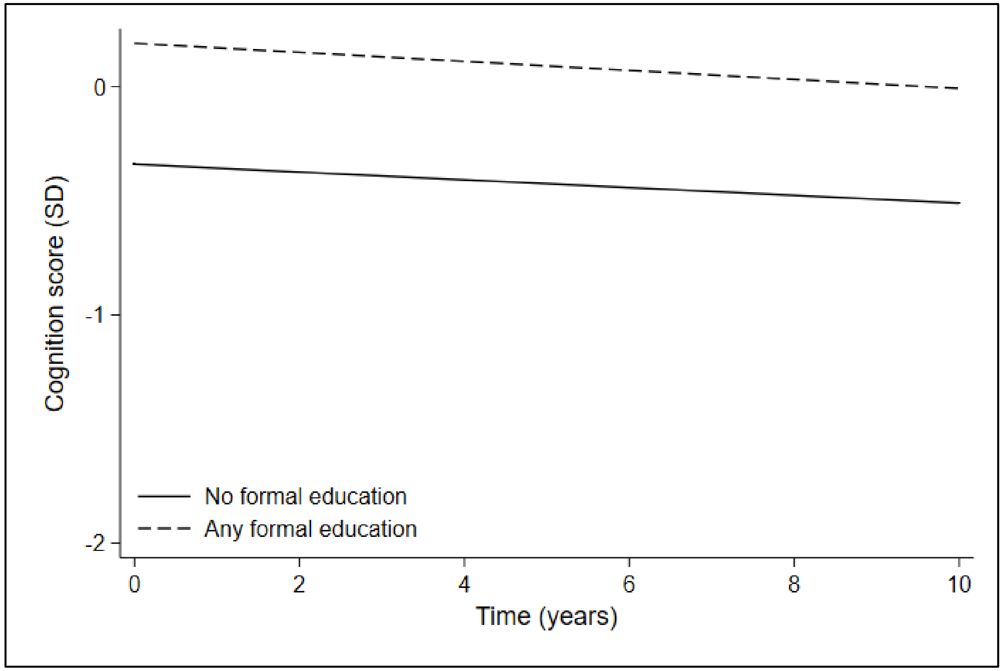

*Figure 1. Cognition scores over time by educational attainment, adjusted for age, sex, socio-economic position, and BMI*

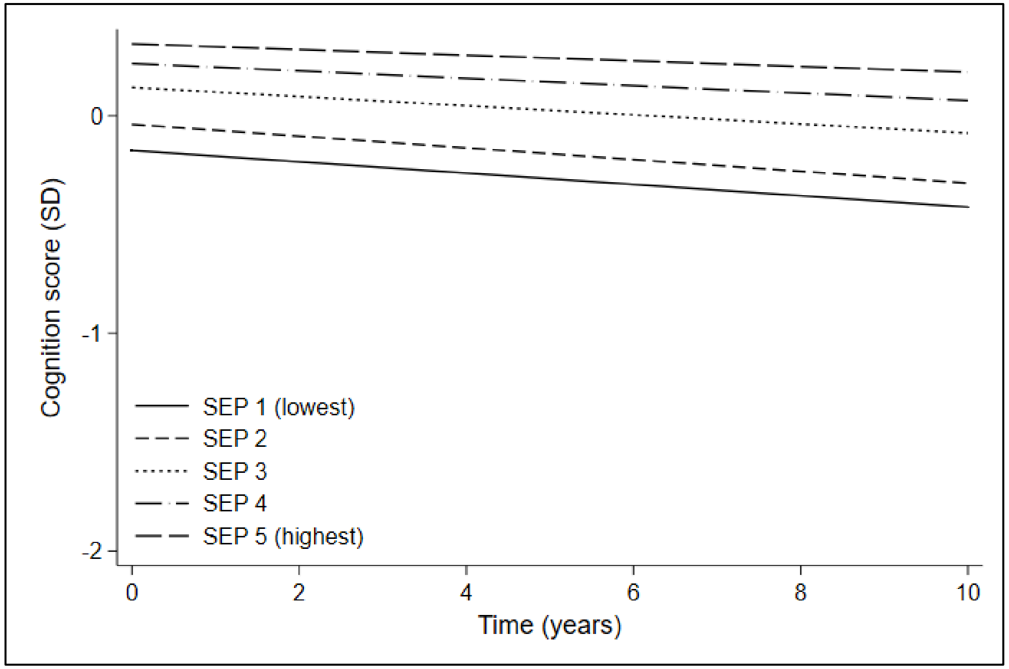

*Figure 2. Cognition scores over time by socio-economic position (SEP), adjusted for age, sex, educational attainment, and BMI*
